# Long-term Durable Humoral Immune Response to Heterologous Antigenic Exposure Post six months by Natural SARS-CoV-2 Infection and Vaccination

**DOI:** 10.1101/2022.02.23.22271381

**Authors:** Gaurav Batra, Deepika Rathna Murugesan, Souvick Chattopadhyay, Farha Mehdi, Ayushi, Mudita Gosain, Savita Singh, Soon Jyoti Das, Suprit Deshpande, Jayanta Bhattacharya, Anil K Pandey, Sankalp Jha, Shweta Goswami, Asim Das, Tanima Dwivedi, Nandini Sharma, Suresh Kumar, Pragya Sharma, Seema Kapoor, Pallavi Kshetrapal, Nitya Wadhwa, Ramachandran Thiruvengadam, Rakesh Kumar, Ritu Gupta, Pramod Kumar Garg, Shinjini Bhatnagar, DBT Consortium for COVID-19 Research

## Abstract

**Importance:** Both vaccination and natural infection lead to immunity and may augment mutual immune response against SARS-CoV-2. There is a need for an evidence-driven booster vaccination policy depending on durability of immune response.

**Objective:** To determine the durability of humoral immune response with varying age, vaccine type, duration, and previous natural infection at least six months after complete vaccination with ChAdOx1 nCov-19 or BBV152.

**Design:** Cross-sectional observational study conducted between November 2021 and January 2022.

**Setting:** Participants were drawn from a DBT COVID-19 Research Consortium cohort in Delhi National Capital Region, India.

**Participants:** We included 2003 individuals who had completed six months after complete vaccination: (i) vaccination with ChAdOx1 nCoV-19 and aged 18-59 years, (ii) vaccination with ChAdOx1 nCoV-19 and aged ≥60 years (iii) vaccination with BBV152 and aged 18-59 years (iv) vaccination with BBV152 and aged ≥60 years (v) vaccination with either vaccine plus SARS-CoV-2 infection referred as those having hybrid immunity. A group of 94 unvaccinated individuals was also included for comparison.

**Exposure:** Age, vaccination type, prior SARS-CoV-2 infection and duration from vaccination/infection.

**Main Outcome(s) and Measure(s):** Humoral immune response determined by anti-RBD IgG concentrations and the presence of anti-nucleocapsid IgG.

**Results:** The serum anti-RBD IgG antibodies were detected (cut-off 24 BAU/ml) in 85% participants with a median titer of 163 (IQR 73, 403) BAU/ml. In the hybrid immunity group, 97.6% [295 (IQR 128, 687) BAU/mL] tested positive for anti-RBD IgG compared to 81.3% [139 (IQR 62, 326) BAU/ml] of only vaccinated participants [χ2 test: p <0.001]. The median anti-RBD IgG titers were higher in the ChAdOx1 nCoV-19 versus BBV152 groups. The median anti-RBD IgG titer in the anti-nucleocapsid positive participants [326 (IQR 132, 739) BAU/ml] was significantly higher than in those without anti-nucleocapsid antibodies [131 (IQR 58, 288) BAU/ml; p <0.001]. The IgG anti-RBD antibodies was present in 85% of participants beyond a median of 8 months after complete vaccination.

**Conclusions and Relevance:** Considering the wide seropositivity rates due to natural SARS-CoV-2 infection, recommendation for boosters should take into account past infections in the population.

**Key points:** *Question:* What is the extent of waning of humoral immune response in various groups of vaccinated individuals at least six months after complete vaccination with ChAdOx1 nCov-19 or BBV152 with or without prior natural infection?

*Findings:* Cross-sectional observational study demonstrates persistence of anti-RBD IgG in 85% of participants even beyond a median of 8 months after complete vaccination. The antibody concentrations were significantly higher in those with hybrid immunity.

*Meaning:* Humoral immunity may last longer due to heterologous antigenic exposure following vaccination and natural infection emphasizing the need for contextualizing the booster policy.

---

Vaccination is the most effective preventive strategy against SARS-CoV-2. Vaccines designed against the ancestral virus act primarily by inducing neutralizing antibodies (Nabs) against the receptor-binding domain (RBD) of viral spike (S) protein. Multiple waves of virus infection led by variants of concern (VoCs) such as alpha (B.1.1.17), delta (B.1.617.2), and omicron (B.1.1.529) have led to breakthrough infections. Others and we have shown that antibody levels wane over time.^1–3^ Antigenic exposure through both vaccination and natural infection may act synergistically to boost immune responses.^4^ There is a need for an evidence-driven booster vaccination policy depending on durability of immune response. Vaccination against SARS-CoV-2 was started in January 2021 in India using an adenovirus vectored recombinant coronavirus vaccine, ChAdOx1 nCov-19 (Covishield, Serum Institute of India), and a whole virion inactivated SARS-CoV-2 vaccine, BBV152 (Covaxin, Bharat Biotech). We hypothesized that the durability of immune response would vary with age, vaccine type, duration, and previous natural infection. Hence, we studied the humoral immune responses in various groups of vaccinated individuals, with or without prior SARS-CoV-2 infection at least six months after complete vaccination.

## Methods

The participants were sampled cross-sectionally from an ongoing DBT COVID-19 Research Consortium cohort between November 2021 and January 2022 (Appendix). We included five groups of individuals who had completed six months after complete vaccination: (i) vaccination with ChAdOx1 nCoV-19 and aged 18-59 years, (ii) vaccination with ChAdOx1 nCoV-19 and aged ≥60 years (iii) vaccination with BBV152 and aged 18-59 years (iv) vaccination with BBV152 and aged ≥60 years (v) vaccination with either vaccine plus prior SARS-CoV-2 infection referred as those having hybrid immunity. A group of unvaccinated individuals was also included for comparison. Written informed consent was obtained from the study participants. Institute ethics committees approved the study.

The study participants were interviewed for details on SARS-CoV-2 vaccination including the type, number of doses, and date of vaccination, profession, comorbidities and prior SARS-CoV-2 infection. The dates of vaccination were verified by the certificate issued by Government of India. Five percent of interviews were monitored for quality control.

Anti-RBD IgG concentrations in the plasma samples were determined by enzyme-linked immunosorbent assay (ELISA) as described earlier with minor modifications and are reported as BAU/mL.^5^ In addition, anti-nucleocapsid antibodies were detected using a qualitative ELISA.

We estimated a sample size of at least 296 participants for each group to detect a decline of anti-IgG RBD by 30% after 6 months of vaccination with 80% power and at 5% significance after adjusting for multiple comparisons. The baseline characteristics are given as median (interquartile range) or number (percentage) as appropriate. The differences in ELISA titers between different groups were compared using unpaired Student’s t-test after log2 transformation. Categorical parameters were compared by chi-square test. All statistical analysis was done using R 4.1.1. A p-value of <0.05 was considered statistically significant.

## Results

We included 2003 vaccinated (1005 ChAdOx1 nCoV-19 and 998 BBV152) and 94 unvaccinated individuals(Figure S1). The median duration after the second dose of vaccine to inclusion was 246 days (IQR: 222, 283). Prior SARS-CoV-2 infection was reported by 21% (421 of 2003) vaccinated and 87% (82 of 94) unvaccinated individuals. The characteristics of participants are provided in Table 1.

**Table-1.** Clinical characteristics of participants stratified by participant groups.

| **Characteristic**<br>**N (%)** | **ChAdOx1**<br>**nCoV-19,**<br>**<60y,**<br>**uninfected,**<br>**N = 388** | **ChAdOx1**<br>**nCoV-19,**<br>**≥60y,**<br>**uninfected,**<br>**N = 352** | **BBV152,**<br>**<60y,**<br>**uninfected,**<br>**N = 465** | **BBV152,**<br>**≥60y,**<br>**uninfected,**<br>**N = 377** | **ChAdOx1**<br>**nCoV-19 /**<br>**BBV152 and**<br>**infected,**<br>**N = 421** | **Unvaccinated,**<br>**N = 94** |
| --- | --- | --- | --- | --- | --- | --- |
| Age (years)* | 34 (27, 46) | 67 (63, 71) | 43 (34, 52) | 66 (63, 70) | 50 (37, 62) | 34 (25, 48) |
| Female | 130 (34%) | 141 (40%) | 165 (35%) | 169 (45%) | 133 (32%) | 30 (32%) |
| Comorbidity | 30 (7.7%) | 107 (30%) | 40 (8.6%) | 105 (28%) | 77 (18%) | 0 (0%) |
| Frontline worker # | 195 (50%) | 21 (6.0 %) | 104 (22%) | 8 (2.1%) | 112 (27%) | 1 (1.1%) |
| Duration between<br>two doses (days)* | 35 (30, 49) | 43 (33, 51) | 31 (30, 36) | 33 (30, 41) | 36 (31, 50) | NA |
| Duration after<br>vaccination * | 274 (237,<br>301) | 230 (216,<br>243) | 265 (226,<br>305) | 246 (232,<br>262) | 241 (206,<br>277) | NA |

\*median (Interquartile range)

The anti-RBD IgG antibodies were undetectable (cut-off of 24 BAU/ml) in 15% (300 of 2003) of the vaccinated participants. The median serum anti-RBD IgG antibody titer among the ELISA positive participants was 163 (IQR 73, 403) BAU/ml. In the hybrid immunity group, 98% tested positive for anti-RBD IgG compared to 81% of only vaccinated participants (p <0.001). The anti-RBD IgG titer in those with hybrid immunity [295 (IQR 128, 687)] BAU/mL was significantly higher than only vaccinated participants [139 (IQR 62, 326); p <0.001]. The median anti-RBD IgG titers were higher in the ChAdOx1 nCoV-19 groups than in the BBV152 groups. In the ChAdOx1 nCoV-19 group, the median anti-RBD IgG titers were higher in ≥60 years participants [242 (IQR 106, 583) BAU/ml] compared to those <60 years [150 (IQR 71, 314) BAU/ml; p <0.001] (Table 2, Figure 1).

**Table 2.**
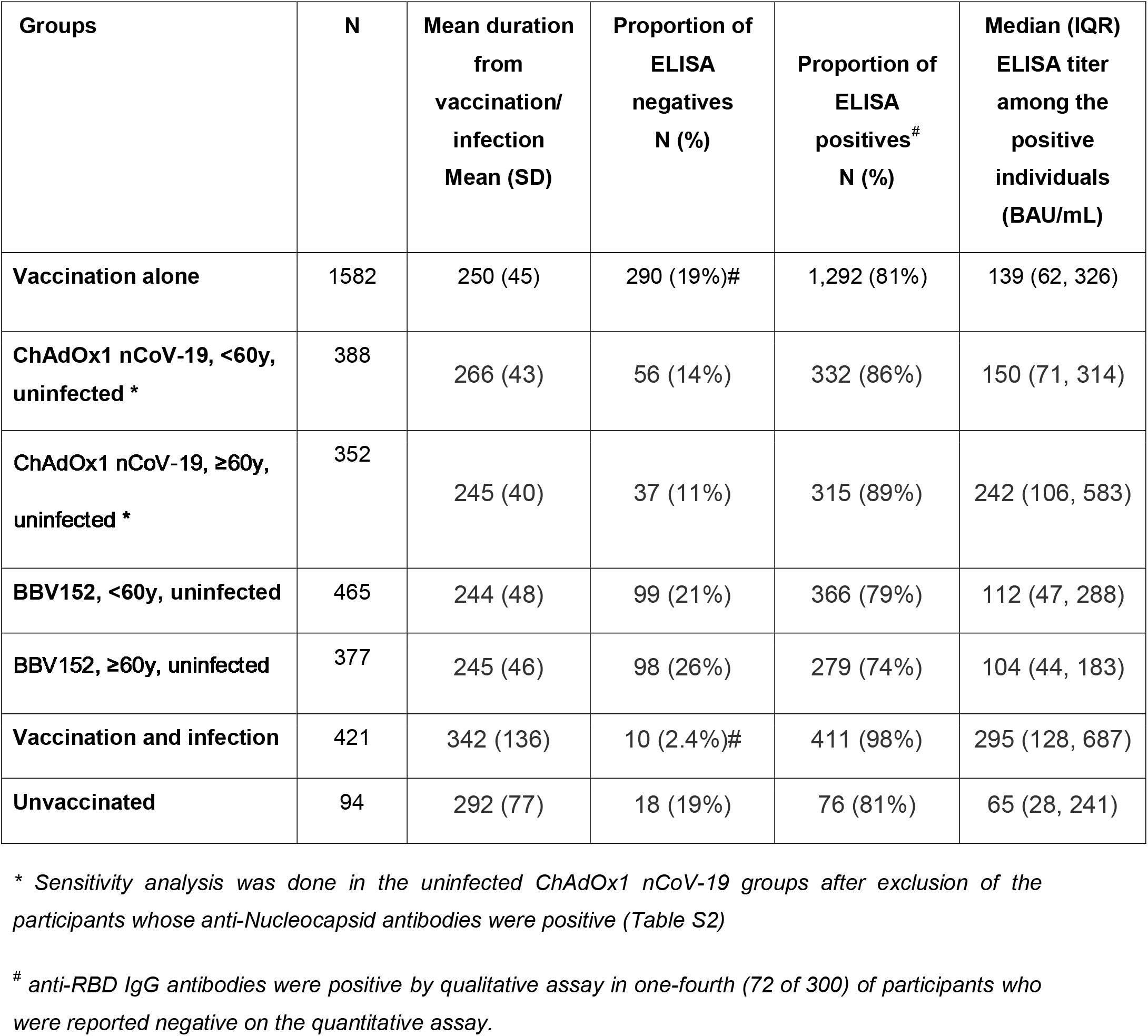
Anti-RBD IgG titers (BAU/mL) among different groups of participants.

| **Groups** | **N** | **Mean duration**<br>**from**<br>**vaccination/**<br>**infection**<br>**Mean (SD)** | **Proportion of**<br>**ELISA**<br>**negatives**<br>**N (%)** | **Proportion of**<br>**ELISA**<br>**positives<sup>#</sup>**<br>**N (%)** | **Median (IQR)**<br>**ELISA titer**<br>**among the**<br>**positive**<br>**individuals**<br>**(BAU/mL)** |
| --- | --- | --- | --- | --- | --- |
| **Vaccination alone** | 1582 | 250 (45) | 290 (19%)<sup>#</sup> | 1,292 (81%) | 139 (62, 326) |
| **ChAdOx1 nCoV-19, <60y,**<br>**uninfected \*** | 388 | 266 (43) | 56 (14%) | 332 (86%) | 150 (71, 314) |
| **ChAdOx1 nCoV-19, ≥60y,**<br>**uninfected \*** | 352 | 245 (40) | 37 (11%) | 315 (89%) | 242 (106, 583) |
| **BBV152, <60y, uninfected** | 465 | 244 (48) | 99 (21%) | 366 (79%) | 112 (47, 288) |
| **BBV152, ≥60y, uninfected** | 377 | 245 (46) | 98 (26%) | 279 (74%) | 104 (44, 183) |
| **Vaccination and infection** | 421 | 342 (136) | 10 (2.4%)<sup>#</sup> | 411 (98%) | 295 (128, 687) |
| **Unvaccinated** | 94 | 292 (77) | 18 (19%) | 76 (81%) | 65 (28, 241) |

*\* Sensitivity analysis was done in the uninfected ChAdOx1 nCoV-19 groups after exclusion of the participants whose anti-Nucleocapsid antibodies were positive (Table S2)*
*<sup>#</sup> anti-RBD IgG antibodies were positive by qualitative assay in one-fourth (72 of 300) of participants who were reported negative on the quantitative assay.*

**Figure 1:**
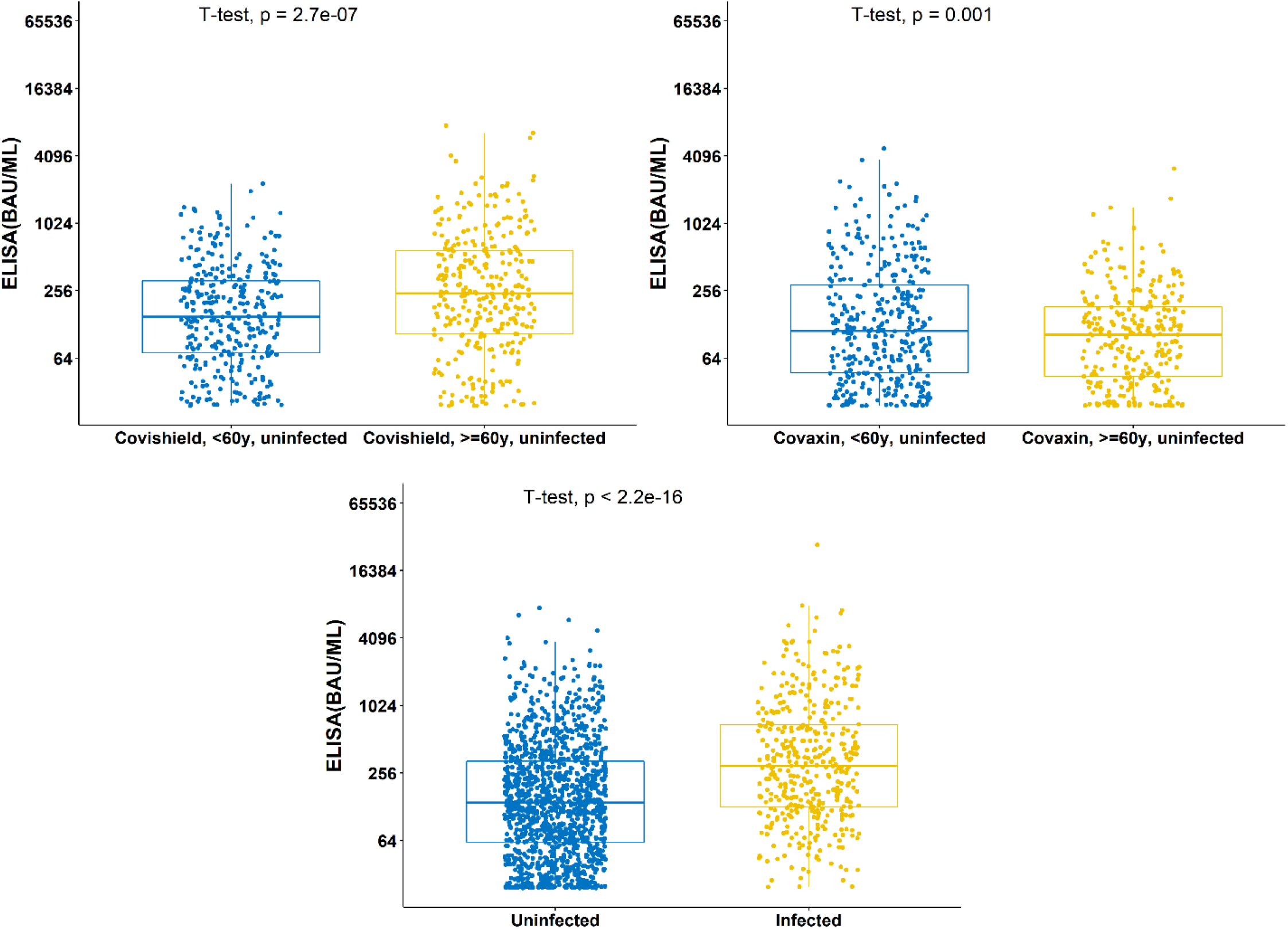
Comparison of anti-RBD IgG titer (BAU/mL) in different participant groups

Anti-nucleocapsid antibodies, which may be induced by natural infection or the whole inactivated virus BBV152 vaccine, were detected in one-fourth (505 out of 2003) of the vaccinated population. The prevalence of anti-N antibodies was 36% (152 of 421) in the hybrid immunity group and 28% (237 of 842) in the uninfected BBV152 vaccinated individuals. The prevalence in the ChAdOx1 nCoV-19 group was 9.5% (37 of 388) in those <60 years and 22% (79 of 352) in those aged 60 and above. The median anti-RBD IgG titer in the anti-nucleocapsid positive participants [326 (IQR 132, 739) BAU/ml] was significantly higher than those without anti-nucleocapsid antibodies [131 (IQR 58, 288) BAU/ml; p <0.001] (Table S1).

## Discussion

In summary, we have shown the durability of IgG anti-RBD antibodies in 85% of participants beyond a median of 8 months after complete vaccination. The antibody concentrations were significantly higher in those with hybrid immunity.

Humoral immune responses to mRNA vaccines are known to wane over a period of time, particularly after 5 months, more so in individuals older than 60 years of age.^6,7^ But these may not be generalizable to other vaccine types and in different geographic regions in the context of immunity provided by natural infection. In the present study, we found higher positivity and higher levels of antibodies among those with hybrid immunity. In the vaccinated group, it is probable that many participants had prior asymptomatic infection consistent with a high seroprevalence of natural infection in the population.^8,9^ We found 15% ChAdOx1-nCoV-19 only participants positive for anti-nucleocapsid antibody suggesting previous infection. However, in the vaccination plus infection group, nearly two-thirds of participants were negative for anti-nucleocapsid antibody possibly suggesting faster waning of anti-nucleocapsid than anti-RBD antibodies.^10^ If we extrapolate this finding to vaccine alone groups, 45% ChAdOxnCoV-19 group might have had asymptomatic natural infection and a similar percentage in the BBV-152 vaccine given the almost similar vaccine effectiveness of these 2 vaccines.^11,12^

This inference is important because most countries have been affected by a minimum of three waves of different SARS-CoV-2 VoCs, most recent being omicron, infecting a large proportion of population. Such a natural infection should be equivalent to at least another dose of vaccine, reinforcing the protection provided by vaccination alone. It is likely that other modes of protection such as cellular responses provided by natural infection are better than those induced by vaccines. Clinically, naturally infected people have shown to be better protected than vaccination alone.^13^ Current data suggest that vaccination plus natural infection provides even better protection.^14,15^ Therefore, recommendation for boosters should take into account past infections in the population.

The strengths of our study include inclusion of inactivated vaccine about which sparse data are available, adequate sample size, inclusion of different age groups and profiling anti-nucleocapsid antibody positivity for unidentified past infections. Our study has a few limitations, inability to unequivocally identify individuals with prior infection, and a small sample size of unvaccinated group, a reflection of >90% of eligible population having been vaccinated in India as in many other countries.

Our data may guide policy for a booster dose. Considering the wide seropositivity rates due to natural infection, the priority for a booster should be for vulnerable and high-risk groups.

## Data Availability

The deidentified dataset and related codes for analysis may be available to researchers upon request addressed to the corresponding author after publication after the approval of the Department of Biotechnology, Government of India.

## Acknowledgments

The authors gratefully acknowledge the DBT Consortium for COVID-19 Research for enabling this study. We sincerely thank participants of the study for providing the blood samples.

This research was supported by Ind-CEPI grant (102/IFD/SAN/5477/2018-2019) to Translational Health Science and Technology Institute, Faridabad; COVID-19 Bioresource grant (BT/PR40401/BIOBANK/03/2020); SARS-CoV-2 diagnostics development grant (Grant No. BT/PR40333/COD/139/11/2020) to Translational Health Science and Technology Institute, Faridabad by the Department of Biotechnology and; GIISER South Asia grant from Bill and Melinda Gates Foundation, Seattle, USA.

## Contributors

PKG conceptualized and designed the overall study. SB, RT and NW designed the cohort study; DM, RT, SS, MG, PK, SJ, SG, AD, AP, TD, RG, RK, PS, SK, SKp, NS, SD and JB enrolled the participants, collected the clinical information and pre-processed the biospecimens; SC, FM, SJD and GB developed and performed the ELISA for antibody titres; RT, DM, A and GB managed and analyzed the clinical and laboratory data; PKG, SB, GB, RT reviewed and interpreted the data, and wrote the manuscript; all authors reviewed and approved the final manuscript.

None of the authors were paid by any pharmaceutical company or any other agency to write this article. All authors had full access to the full data in the study and accept responsibility to submit for publication.

## Appendix

### Supplementary Methods

The Department of Biotechnology consortium for COVID-19 Research cohort was developed by Translational Health Science and Technology Institute, Faridabad, India in collaboration with hospitals in Delhi National Capital Region, India, particularly, Employee State Insurance Corporation Medical College and Hospital, Faridabad; Maulana Azad Medical College and associated Loknayak Hospital, New Delhi; and All India Institute of Medical Sciences, New Delhi

#### Assays

Internal positive control was calibrated against the first WHO international standard for anti-SARS-CoV-2 immunoglobulin (code 20/136). (WHO manual for the establishment of national and other secondary standards for antibodies against infectious agents focusing on SARS-CoV-2; Draft Version 12/10/2021). Anti-RBD IgG concentrations above the assay cut-off and corresponding to the linear part of the curve was considered, and values in BAU/ml were assigned to each test sample. Anti-RBD IgG concentrations in the test samples were calculated for each sample dilution by interpolation of OD values on the 4-parameter logistic (4-PL) standard curve from internal positive control using GraphPad Prism 9.3.1 software. Additional dilutions beyond 1:12150 were done for samples where OD values were beyond the calibration curve’s linear part. The RBD and N protein used in ELISA were from the ancestral strain of the virus. The lower limit of quantitation (LLOQ) for the anti-RBD IgG quantitative assay was 24 BAU/ml. For samples below the LLOQ, the presence of anti-RBD IgG was detected using a qualitative ELISA.

**Figure S1:**
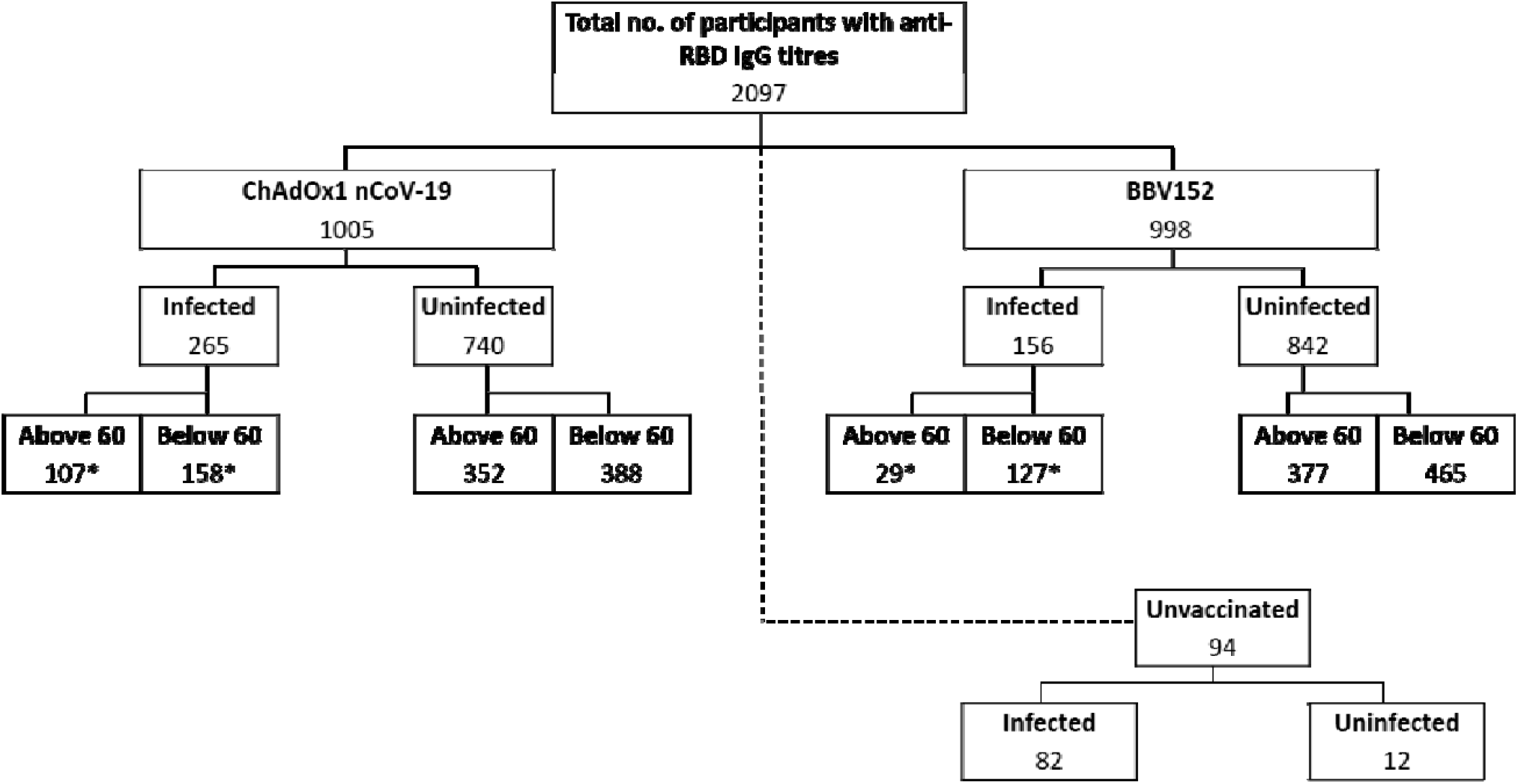
Distribution of study participants among different groups *\*The groups were combined into a group of “vaccinated plus infected individuals”*

**Table S1:** Distribution of anti-nucleocapsid ELISA among different groups of participants.

| **Groups** | **N** | **Anti-N**<br>**Negative** | **Anti-N**<br>**Positive** |
| --- | --- | --- | --- |
| ChAdOx1 nCoV-19, <60y, uninfected | 388 | 351 (90%) | 37 (9.5%) |
| ChAdOx1 nCoV-19, ≥60y, uninfected | 352 | 273 (78%) | 79 (22%) |
| BBV152, <60y, uninfected | 465 | 323 (69%) | 142 (31%) |
| BBV152, ≥60y, uninfected | 377 | 282 (75%) | 95 (25%) |
| ChAdOx1 nCoV-19 / BBV152 + infected | 421 | 269 (64%) | 152 (36%) |
| Unvaccinated | 94 | 71 (76%) | 23(24%) |
| Anti-RBD ELISA negatives | 300 | 289 (19%) | 11 (2.2%) |
| Anti-RBD ELISA positives | 1,703 | 1,209 (81%) | 494 (98%) |
| ELISA titer among the positive<br>individuals<br>(BAU/mL)* | 1,703 | 131 (58, 288) | 326 (132,<br>739) |

\*median (Interquartile range)

**Table S2:** **Anti-RBD IgG titers (BAU/mL) among different groups of participants after exclusion of anti-nucleocapsid positive participants from the ChAdOx1 nCoV-19 only group**

| **Groups** | **N** | **Mean duration from vaccination/ infection Mean(SD)** | **Proportion of ELISA negatives N (%)** | **Proportion of ELISA positives\* N (%)** | **Median (IQR) ELISA titre among the positive individuals (BAU/mL)** |
| --- | --- | --- | --- | --- | --- |
| **Vaccination alone** | 1582 | 250 (45) | 290(19%) | 1,292 (81%) | 139 (62, 326) |
| **ChAdOx1 nCoV-19, <60y, uninfected** | 351 | 267 (43) | 55 (16%) | 296 (84%) | 139 (67, 266) |
| **ChAdOx1 nCoV-19, ≥60y, uninfected** | 273 | 245 (42) | 34 (12%) | 239 (88%) | 213 (95, 464) |
| **BBV152, <60y, uninfected** | 465 | 244 (48) | 99 (21%) | 366 (79%) | 112 (47, 288) |
| **BBV152, ≥60y, uninfected** | 377 | 245 (46) | 98 (26%) | 279 (74%) | 104 (44, 183) |
| **Vaccination and infection** | 421 | 342 (136) | 10 (2.4%) | 411 (98%) | 295 (128, 687) |
| **Unvaccinated** | 94 | 292 (77) | 18 (19%) | 76 (81%) | 65 (28, 241) |

## References

1. Li D, Luan N, Li J, et al. Waning antibodies from inactivated SARS-CoV-2 vaccination offer protection against infection without antibody-enhanced immunopathology in rhesus macaque pneumonia models. Emerging Microbes & Infections. 2021;10(1):2194–2198. doi:10.1080/22221751.2021.2002670

2. Seow J, Graham C, Merrick B, et al. Longitudinal observation and decline of neutralizing antibody responses in the three months following SARS-CoV-2 infection in humans. Nat Microbiol. 2020;5(12):1598–1607. doi:10.1038/s41564-020-00813-8

3. Thiruvengadam R, Chattopadhyay S, Mehdi F, et al. Longitudinal Serology of SARS-CoV-2-Infected Individuals in India: A Prospective Cohort Study. Am J Trop Med Hyg. Published online May 18, 2021:tpmd210164. doi:10.4269/ajtmh.21-0164

4. Reynolds CJ, Gibbons JM, Pade C, et al. Heterologous infection and vaccination shapes immunity against SARS-CoV-2 variants. Science. 2022;375(6577):183–192. doi:10.1126/science.abm0811

5. Mehdi F, Chattopadhyay S, Thiruvengadam R, et al. Development of a Fast SARS-CoV-2 IgG ELISA, Based on Receptor-Binding Domain, and Its Comparative Evaluation Using Temporally Segregated Samples From RT-PCR Positive Individuals. Front Microbiol. 2020;11:618097. doi:10.3389/fmicb.2020.618097

6. Levin EG, Lustig Y, Cohen C, et al. Waning Immune Humoral Response to BNT162b2 Covid-19 Vaccine over 6 Months. New England Journal of Medicine. 2021;385(24):e84. doi:10.1056/NEJMoa2114583

7. Chemaitelly H, Tang P, Hasan MR, et al. Waning of BNT162b2 Vaccine Protection against SARS-CoV-2 Infection in Qatar. New England Journal of Medicine. 2021;385(24):e83. doi:10.1056/NEJMoa2114114

8. Murhekar MV, Bhatnagar T, Thangaraj JWV, et al. Seroprevalence of IgG antibodies against SARS-CoV-2 among the general population and healthcare workers in India, June-July 2021: A population-based cross-sectional study. PLoS Med. 2021;18(12):e1003877. doi:10.1371/journal.pmed.1003877

9. Misra P, Kant S, Guleria R, Rai SK, Aiims WUS study team of. Serological prevalence of SARS-CoV-2 antibody among children and young age (between age 2-17 years) group in India: An interim result from a large multi-centric population-based seroepidemiological study. Published online June 16, 2021:2021.06.15.21258880. doi:10.1101/2021.06.15.21258880

10. Lumley SF, Wei J, O’Donnell D, et al. The Duration, Dynamics, and Determinants of Severe Acute Respiratory Syndrome Coronavirus 2 (SARS-CoV-2) Antibody Responses in Individual Healthcare Workers. Clin Infect Dis. 2021;73(3):e699–e709. doi:10.1093/cid/ciab004

11. Thiruvengadam R, Awasthi A, Medigeshi G, et al. Effectiveness of ChAdOx1 nCoV-19 vaccine against SARS-CoV-2 infection during the delta (B.1.617.2) variant surge in India: a test-negative, case-control study and a mechanistic study of post-vaccination immune responses. The Lancet Infectious Diseases. Published online November 25, 2021. doi:10.1016/S1473-3099(21)00680-0

12. Desai D, Khan AR, Soneja M, et al. Effectiveness of an inactivated virus-based SARS-CoV-2 vaccine, BBV152, in India: a test-negative, case-control study. Lancet Infect Dis. Published online November 23, 2021:S1473-3099(21)00674-5. doi:10.1016/S1473-3099(21)00674-5

13. Goldberg Y, Mandel M, Woodbridge Y, et al. Protection of previous SARS-CoV-2 infection is similar to that of BNT162b2 vaccine protection: A three-month nationwide experience from Israel. Published online April 24, 2021:2021.04.20.21255670. doi:10.1101/2021.04.20.21255670

14. Hammerman A, Sergienko R, Friger M, et al. Effectiveness of the BNT162b2 Vaccine after Recovery from Covid-19. New England Journal of Medicine. 2022;0(0):null. doi:10.1056/NEJMoa2119497

15. Hall V, Foulkes S, Insalata F, et al. Protection against SARS-CoV-2 after Covid-19 Vaccination and Previous Infection. New England Journal of Medicine. 2022;0(0):null. doi:10.1056/NEJMoa2118691

